# Two-phase Framework Clinical Question-Answering – Autocorrection for Guideline-concordance

**DOI:** 10.1101/2024.11.04.24316718

**Authors:** Amara Tariq, Nathan Yu, Bhavik Patel, Imon Banerjee

## Abstract

Use of large language models for generative tasks in critical domains like medicine is fraught with challenges like hallucination. In the domain of medicine, hallucination may take a unique shape where the LLM-generated language is not inaccurate but the suggested treatment or medication has now been discontinued in a specific context. Reinforcement learning based solutions for building reliable LLM-based frameworks are limited by the fact that the reinforcement is typically focused on only identifying the mistake; correcting the mistake is left up to the primary LLM. We propose an innovative solution where a two-phase question answering framework composed of two LLMs is designed such that one LLM learns to generate answers while the other learns to correct any mistakes in the answer generated by the first model. We experimented with the particular domain of prostate cancer and LLMs designed for various domains and showed that domain-specific LLMs outperform generic or wide-domain LLMs.

## Introduction

While large language models (LLM) have proven their utility as zero-shot and few-shot learners with vast application potential [1-4], their use as language generators or question-answering framework in sensitive domains like medicine and healthcare is fraught with several challenges. LLMs trained on data scraped from the World Wide Web are often unfamiliar with medical jargon. Even LLMs designed for the domain of medicine attempt to cover very vast topics including all clinical specialities in medicine. This may result in limited performance on sensitive clinical tasks for these LLMs. Hallucination is a critical challenge in wide-spread adoption of LLMs for tasks requiring specific and accurate use of language [5-7]. This challenge takes an even more critical form in the domains of medicine and healthcare. LLM based question-answering or conversational frameworks (chatbots) may generate language that is discordant with guidelines set by the clinical specialists such as treatment guidelines for cancer by expert societies like American Cancer Society. The generated statements will be considered wrong if they are compliant with out-of-date medical literature e.g., if medications or treatment regimens that are no longer in use are suggested in the LLM-generated language. Past studies have shown that popular state-of-the-art chatbots generated answers discordant with treatment guidelines about 30% of the times [8].

Researchers have been focused on solving the challenge of hallucination or harmful; content generation through reinforcement learning based ideas. A human-in-the-loop type approach is reinforcement learning through human feedback (RLHF) where models are taught to align themselves with humans through feedback provided by humans. The same idea has been modified to use automated feedback through AI based models, i.e., reinforcement learning with AI feedback (RLAIF) [9-11]. Policy optimization approaches; whether direct or proximal (DPO and PPO); are aimed at building safe and reliable models by designing policies focused around safety and reliability. These approaches have found wide-spread adoption in the domain of LLM based model development [12-14]. Frameworks are usually composed of a primary model (LLM-based task model) and a secondary “ policy” model. Literature survey reveals limitations of these frameworks in terms of computational cost and instability of reinforcement learning [15].

We argue that a critical limitation of these approaches towards reliable and safe LLM based model development is that the policy modules are only aimed at identifying mistakes and not correcting them themselves. The onus of correction is placed on the primary LLM based model itself. This approach is not ideal for medical domain question-answering or chatbot as the notion of “ correct” may evolve over time. A classic example is treatment guidelines set by experts societies like American Cancer Society for different types of cancer, e.g., prostate cancer. As new research leads to development of pharmaceuticals and other treatment regimens, these guidelines are updated. An LLM-based chatbot trained on large amounts of conversational data with optimized policy of correct answer generation will start generating “ incorrect” or guideline-discordant answers after such an update. Policy optimization based LLM frameworks will require policy update and finetuning of the primary LLM to potentially correct these mistakes.

We propose a novel two stage framework for LLM-based question-answering where two LLM-based frameworks; one primary and one secondary; are trained with one focused on answering questions and the other on not only evaluating the answer for correctness but also modifying it to correct the generated statement if needed, i.e., auto-correction. The proposed solution to building a reliable LLM-based framework is different from previously proposed approaches based on reinforcement learning as it breaks down the task in two parts, leaving open the possibility of finetuning one model if needed. We experimented with the use-case of question-answering regarding treatments for prostate cancer such that the correctness criteria was based on compliance with treatment guidelines set by the American Cancer Society. As described earlier, the notion of correctness evolves over time in a domain like medicine. Secondary model in the framework which is focused on correcting the mistakes made by the first model can be updated separately to translate out-of-date content such as a medication that is no longer in use, into latest content such as some newly developed and approved medication or treatment regimen.

## Materials and Methods

The proposed framework as shown in Figure 1 is composed of two models; the primary model is focused on answering the question provided as input (QA model) and the secondary model (translator) is supposed to translate the answers generated by the primary model into statements compliant with the treatment guidelines set by American Cancer Society. The secondary model will only make modifications if the answer generated by the primary model is discordant with the treatment guidelines.

**Figure 1:**
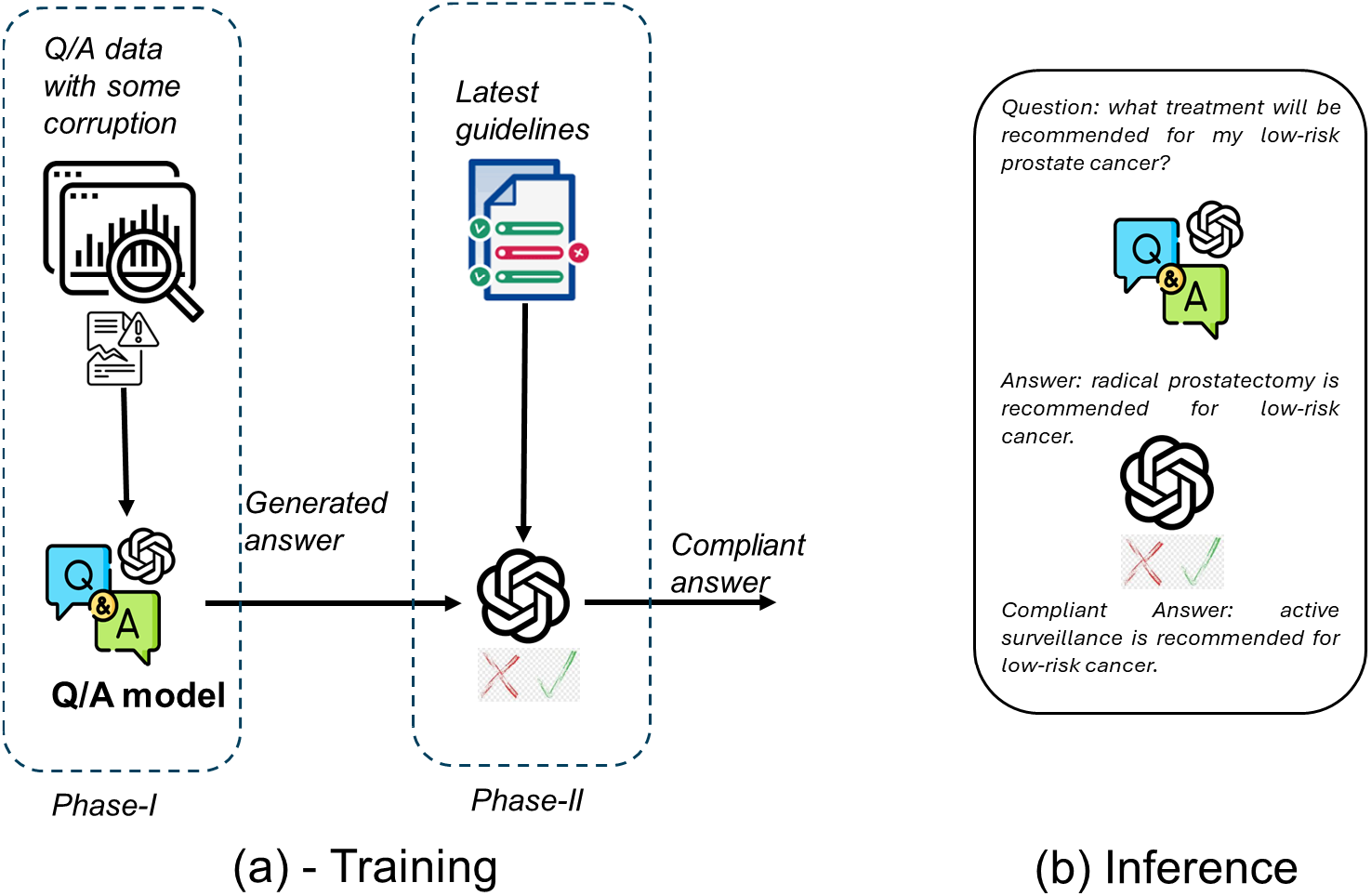
Proposed two-phase question answering framework in training and inference stages; phase-I is designed to answer questions and phase-II is designed to correct any mistakes made in phase-I.

### Dataset

We manually curated 320 questions inspired by all sections of the prostate cancer treatment guidelines set by the American Cancer Society. These guidelines are publicly available online. A board-certified radiation oncologist curated “ non-compliant” or “ corrupt” answers for each question. We then used open-source LLaMA-7B-chat [16] to paraphrase these questions to form an extended dataset. After manual filtering for misleading or incomplete paraphrases, we were left with 780 question-answer-corrupt answer tuples. Approximately 5% tuples were held-out for evaluation purposes.

### Model development

The proposed framework is composed of two generative language models. We experimented with three LLMs as basis for the proposed QA framework; 1-GPT2 [17], 2-BioGPT [18], 3-PCaLLM [19]. All three were similarly sized at around 340M parameters. We aimed at covering the wide landscape of LLMs by including LLMs designed with different scopes and for different domains. GPT2 was trained using web-carawled text and can be considered a general-purpose or generic LLM designed for a wide scope of language. BioGPT was specifically designed for the broad fields of medicine and biology and was trained on the PubMed dataset [18]. In contrast, PCa-LLM was designed specifically for the domain of prostate cancer and was trained on a combination of de-identified clinical notes and radiology and pathology reports collected from a large multi-site healthcare institution as well as prostate cancer related data from PubMed. This model is coupled with a domain-specific tokenizer that tends to cover clinical terminology regarding prostate cancer much better than general purpose LLMs. We employed similarly-sized versions of all models (340M).

### Training simulation

We attempted at simulating a realistic scenario where some treatment regimens and medication become guideline-discordant over time as new guidelines are published. During training, we randomly selected N% questions and replaced their correct answers with corrupt answers. During inference, only compliant answers were used as ground truth. We experimented by varying N between 1 and 10 percent to test the robustness of the framework.

## Results

Table 1 shows performance of the proposed framework with different LLMs in terms of quality of answer. It is clear that the phase-II or the secondary LLM of the proposed framework is effective in mitigating the mistakes made by the phase-I. However, the efficacy of this approach seems dependent on the chosen LLM base model.

**Table 1.** Performance of the proposed framework with different LLM base models.

| Percentage of corrupt QA-pairs | Incorrect answers (out of 25 test set questions) |  |  |  |  |  |
| --- | --- | --- | --- | --- | --- | --- |
|  | GPT2 |  | BioGPT |  | PCaLLM |  |
|  | phase-I | phase-II | phase-I | phase-II | phase-I | phase-II |
| 1% | 10 | 12 | 7 | 8 | 9 | 5 |
| 5% | 7 | 6 | 5 | 6 | 8 | 3 |
| 10% | 15 | 16 | 5 | 0 | 15 | 10 |

**Table 2.** Sample answers generated by the proposed framework.

| Model | Question | What are the options for managing cancer that has disseminated to adjacent organs following surgical removal? |
| --- | --- | --- |
|  | answer | Radiation therapy to the pelvic area might be an option, typically along with hormone therapy (which might include a newer hormone drug such as abiraterone) if cancer is found in nearby organs after surgery |
| GPT2 | Phase-I answer | hormone therapy, monitoring the cancer, surgery to remove the pelvic lymph nodes, or radiation to the pelvic lymph nodes may be used if cancer is found |
|  | **Phase-II answer** | hormone therapy, monitoring the cancer, surgery to remove the pelvic lymph nodes, or radiation to the pelvic lymph nodes may be used if cancer is found |
| **BioGPT** | **Phase-I answer** | radiation therapy to the pelvic area might be an option, typically along with hormone therapy |
|  | **Phase-II answer** | radiation therapy to the pelvic area might be an option, typically along with hormone therapy |
| **PCaLLM** | **Phase-I answer** | Chemotherapy and hormone therapy are not likely to be helpful |
|  | **Phase-II answer** | Radiation therapy along with the hormone therapy is typically used |
|  | **Question** | In what clinical contexts might second-generation anti-androgens be employed to manage prostate cancer? |
|  | **Answer** | Second generation anti androgens like Enzalutamide can be used in case a biochemical recurrence, and that is thought to be at high risk for spreading to other parts of the body |
| **GPT2** | **Phase-I answer** | second generation anti androgens drugs can be helpful in men with cancer that has spread but that is still responding to other forms of hormone therapy |
|  | **Phase-II answer** | Newer generation anti androgens drugs can be helpful in men with cancer that has spread but that is still responding to other forms of hormone therapy |
| **BioGPT** | **Phase-I answer** | Second generation anti androgens drugs can also be combined with orchiectomy or an LHRH agonist as first-line hormone therapy. |
|  | **Phase-II answer** | CAB is a second generation anti androgen drug. This is called combined androgen blockade (CAB). |
| **PCaLLM** | **Phase-I answer** | As men age, prostate cancer often times two or more than men may have higher prostate cancer risk factors for which treatment is typically recommended. |
|  | **Phase-II answer** | The goals of this treatment are to reduce the risk of prostate cancer recurrence, spread to nearby organs, and to improve the quality of life of the patient's |

## Discussion

In this study, we propose a novel and adaptable solution to the challenge of reliable LLM-based framework development for critical and sensitive domains like medicine and healthcare. We chose the use-case of question answering particularly for prostate cancer with reliability criteria set as compliance with evolving treatment guidelines. We included a variety of LLMs in our experiments including generic LLM like GPT-2 trained on vast amounts of web-crawled text, biomedical focused BioGPT trained on research literature, and domain-specific LLM for prostate cancer trained on de-identified textual patients’ data including clinical notes, reports in addition to domain-specific research literature.

The proposed solution is composed of two phases with two generative models; one primary (QA model) and one secondary (compliance translator). The primary model is trained to answer questions and the secondary model is trained to fix mistakes or out-of-compliance treatment suggestions in the answers generated by the first model. Only the second model needs fine tuning when the treatment guidelines are updated.

The results in Table 1 showed the efficacy of our approach. We observed that the framework built on top of our domain-specific LLM (PCaLLM) was consistently able to correct some of the mistakes made by the primary module in the operation of the secondary module. As expected, a larger and larger number of errors were made by the model. We also observed that the PCaLLM based framework easily outperforms the GPT2 based framework. This observation is intuitive as GPT2 is not particularly focused on prostate cancer or even medicine, While GPT2 has been established as a zeroshot framework for a wide variety of problems, there are limits to its application on specialized tasks like QA about prostate cancer. However, our QA dataset was based on treatment guidelines published by the American Cancer Society which are openly available online. Since GPT2 was trained on web-scrapped text, the text of guidelines is likely part of the pretraining data for GPT2. This is not the case for our PCaLLM which was pretrained on clinical notes of prostate cancer patients and prostate cancer related abstracts from PubMed dataset. Still, domain-focused pretraining enabled PCaLLM-based framework to perform better that GPT2-based framework.

Medicine-focused BioGPT was pre trained on all PubMed data. BioGPT-based QA framework demonstrated an interesting pattern. While its primary module tended to make fewer mistakes than PCaLLM based framework’s primary module, it was unable to correct these mistakes during the operation of the secondary module unless the amount of corruption was increased to 10%. We believe that this was due to the overfitting of the module to the small training set. During the training of the secondary module, the module was optimized to only change the incorrect/corrupt/discordant answers which represented a small portion of the training data (1%-10%). In the BioBGPT based framework, the secondary module seemed to stick to the behavior of not altering its input until its training data was changed to include a significant portion of samples where input text needed alteration to match the groundtruth.

Our study represents an innovative generative solution to the issue of reliable text generation in a sensitive domain like medicine where the nation of correctness evolves over time as new research leads to new treatments, regimens and medications. The task of answering and correcting the answer to enhance reliability is split into two modules such that only the secondary module will need to be retrained to keep up with the evolving clinical research. In the future, we plan to evaluate our framework on a large-scale data of patient-provider conversation.

## Data Availability

All data produced in the present study are available upon reasonable request to the authors.

## Data availability statement

Prostate cancer treatment guidelines set by the American Cancer Society are available at https://www.cancer.org/cancer/types/prostate-cancer/treating.html. Curated question-answer pairs using the text of treatment guidelines are available upon reasonable request to the authors.

